# Response to neoadjuvant chemoradiotherapy in rectal cancer is associated with RAS/AKT pathway dysregulation and high tumour mutational burden

**DOI:** 10.1101/2020.02.01.20019794

**Authors:** Joanne D Stockton, Louise Tee, Celina Whalley, Jonathan James, Mark Dilworth, Rachel Wheat, Thomas Nieto, Ian Geh, Andrew D Beggs

**Author notes:** Correspondence to: Andrew Beggs, Surgical Research Laboratory, Institute of Cancer & Genomic Science, University of Birmingham, Vincent Drive, Birmingham, B15 2TT. Contributorship: Manuscript: ADB, JS, IG, DM; Molecular analysis: JS, LT, CW, JJ, MD, TN, AB; Sample acquisition and patient cohort: IG, DM, ADB. Statistical analysis: Andrew Beggs. Data sharing statement: All sequence data will be uploaded to the SRA archive on publication of this study. All methylation microarray data will be uploaded to the GEO archive on publication of this study.

## Abstract

**Purpose:** Pathological complete response (pathCR) in rectal cancer, seen in examination of the pathological specimen post-surgery is the phenomenon whereby a tumour completely regresses under treatment with chemoradiotherapy. This is beneficial as up to 75% of patients do not experience regrowth of the primary tumour, allowing organ preservation and is poorly understood. We aimed to characterise the processes involved in pathCR.

**Materials & Methods:** Two groups of patients were identified with either complete response (pathCR group) or no response (poor response group) and biopsy and/or resection specimen blocks were retrieved. These underwent high read depth amplicon sequencing, exome sequencing, methylation arrays and immunohistochemistry for DNA repair pathway proteins. Sequencing data underwent analysis and the two cohorts were compared.

**Results:** Seven patients who underwent pathological complete response and twenty four who underwent poor response (to act as opposite “extreme phenotypes”) underwent molecular characterisation. Patients in the complete response group had significantly higher tumour mutational burden, neoantigen load and enrichments for mutations in the PI3K/AKT/mTOR signalling pathway as well as significantly lower numbers of structural variants. There were no differences in copy number variants or tumour heterogeneity. Methylation analysis demonstrated enrichment for changes in the PI3K/AKT/mTOR signalling pathway.

**Conclusions:** The phenomenon of pathCR in rectal cancer appears to be related to immunovisibility caused by a high tumour mutational burden phenotype. Resistance mechanisms seem to involve the PI3K/AKT/mTOR signalling pathway and tumour heterogeneity does not seem to play a role in resistance.

## INTRODUCTION

Rectal cancer is a common malignancy (1), with approximately 11,000 case per year in the UK(2). Treatment typically consists of excisional surgery (3) with neoadjuvant therapy if the cancer is locally advanced, consisting of either short course radiotherapy (4) (25 Gy in 5 fractions over 1 week) with surgery on the following week, or long course neo-adjuvant chemoradiotherapy (nCRT) (5) (45-50 Gy in 25 fractions over 5 weeks, with synchronous iv 5-FU or oral capecitabine) with surgery 6-10 weeks later. This latter regimen leads to significant tumour shrinkage and downstaging, with pathological complete response (pathCR, defined as complete regression of tumour in the resection specimen) observed in approximately 10-15% of patients (6). Multiple investigators have shown higher rates of response (7) (8) with higher radiotherapy doses, with pCR rates of 25-30%.

Clinical complete response (CCR) is defined as the absence of tumour on imaging and/or clinical examination, but does not definitely exclude residual tumour, which requires a resection specimen to confirm. If high rates of CCR can be associated with pathCR, this may allow the use of watch and rate to occur routinely. Pathological CR is associated with several potential benefits., either effecting potential cure (9) (defined as >5 years recurrence free) or either delaying the need for excisional surgery (10) or allowing potential organ preservation surgery (11) such as TEMS or TAMIS (12). It is unclear as to what the molecular drivers of pathCR are (13, 14), but they are thought to relate to the factors that promote radiotherapy and chemotherapy related tumour killing. Rectal cancers exist within a low oxygen tension environment (15) leading to an intrinsic resistance. As radiotherapy induces the formation of oxygen derived free radicals, which cause tumour cell death by direct DNA damage, a low oxygen environment leads to fewer available oxygen free radicals, leading to lower cell death.

Another hypothesised mechanism is around DNA repair. Ionising radiation (16) induces a DNA double strand break (DSB), which is then repaired either by one of two mechanisms, homologous recombination (HR, where a sister chromatid is used to repair the defect) or non-homologous end joining (NHEJ), where a complex repair mechanism including BRCA, ATM, ERCC5 and others contribute to a DNA repair complex that re-joins the damaged segments of DNA. There is evidence from previous studies that there is aberrant functioning of the NHEJ pathway is associated with response in radiotherapy (17).

Tumour heterogeneity undoubtedly plays a role (13, 18), as does immunogenicity caused by the formation of clonal neoantigens which determines the immune microenvironment. As the rates of microsatellite instability in rectal cancer are low (2-3% (19)), there is little hypermutation & therefore rates of immunovisibility in rectal cancer are low. However Akiyoshi et al (1) have recently shown that levels of clonal neoantigens are higher in patients undergoing a good response to neoadjuvant chemoradiotherapy, suggesting that hypermutation plays a role as this is the primary mechanism by which neoantigens are produced. A previous integrated molecular analysis of rectal cancer (13) focusing on resistance to rectal cancer found no key genomic features that correlated with resistance, in opposition to that found in Akiyoshi’s study.

Because of the uncertainty surrounding the precise mechanisms of sensitivity of rectal cancer to chemoradiotherapy, we aimed to study the phenomenon of pathCR in rectal cancer by carrying out a comprehensive molecular analysis of both pre-treatment and post-treatment rectal cancer specimens in order to elucidate potential mechanisms of sensitivity.

## MATERIALS & METHODS

### Patients

A prospective database of all patients undergoing neoadjuvant chemoradiotherapy for rectal cancer was used to identify patients. Ethical approval was obtained from the NW Research Ethics committee (15/NW/0079) Patients who underwent long course chemoradiotherapy and achieved pathCR were identified as determined by complete regression of the tumour on histopathological examination of the specimen by a Consultant Histopathologist with absolutely no tumour cells remaining (Mandard grade 1), as opposed to minimal residual disease where several cells were allowable. The histopathology archives were then searched to find the pre-treatment endoscopic biopsies of these patients for downstream analysis. This was defined as the “pathCR cohort”. A randomly selected second cohort of patients were identified, who had poor response to treatment or progressed whilst on treatment, as defined both by post-treatment MRI and Mandard regression grading of either grade 4 or grade 5. This was defined as the “non-responder cohort”. Representative normal tissue from the proximal resection margin was obtained for genomic analysis.

### Samples

Formalin fixed blocks were retrieved for these patients and cut into 4uM sections on frosted glass slides for needle macrodissection and immunohistochemistry. A representative H&E section was used to target tumour cells on needle macrodissection, which was then used to maximise tumour content for DNA extraction. DNA extraction was performed using a modified protocol of the Qiagen DNEasy kit (Qiagen) and eluted DNA was quantified using Nanodrop spectrophotometry (for contaminants) and Qubit fluorimetry (for concentration).

### Immunohistochemistry (IHC)

This was performed on 4uM slides as previously described using a Leica BondMax autostainer. IHC was performed against yH2AX (ab26350), ATM (ab36810), Ku70/80 (ab53126), MLH1 (ab92312), MSH2 (ab52266) and MSH6 (ab14204) and slides were then scanned onto a Leica slide imaging platform. IHC was scored by the QuPath system (20). Briefly, a single section was zoomed to 10x view of a representative area of epithelium/tumour, calibration was performed, either nuclei or membranous counting was set depending on the antibody and auto counting of DAB stained positive/negative cells was carried out. Staining was reported as a percentage of positive/negative stained cells.

### Genomics

Extracted genomic DNA was used for downstream analysis. Targeted amplicon resequencing was performed using the Fluidigm 48×48 Access array. PCR amplicons covering APC, KRAS, BRAF, NRAS, FBXW7, SOX7 (primer sequences available on request) were designed using Primer3 (21). Primer specificity was checked using PrimerBLAST and UCSC in-silico PCR. Briefly, 20ng of FFPE DNA was injected into the Access Array system and thermal cycled according to manufacturer’s specifications. Amplicons were then ligated to Illumina sequencing indexes & adapters and pooled and sequenced on an Illumina MiSeq to an average read depth of > 1000X using a 100bp PE sequencing strategy. For FFPE exome sequencing, a custom modification of the Illumina TruSeq Exome hybridisation kit was used. Approximately 300ng of FFPE-derived DNA was prepared with the following modifications: Firstly, no size selection was performed after end repair and DNA fragments were amplified with 12 cycles of PCR. Enrichment was performed using a bead ratio of 0.8, then samples were combined into pools of 3 plex for coding exome (TruSeq exome, 45mb in size) probe hybridisation and subsequent clean up. 10 cycles of amplification were performed to enrich the final libraries which were then pooled into a final 12 plex library. Sequencing was performed on an Illumina NextSeq 550 75 cycles paired end reads high output flow cell… Methylation was interrogated using the Illumina HumanMethylation 450 array system. Between 100-250ng of FFPE DNA was processed using the Illumina FFPE restore kit, and then hybridised to the HumanMethylation 450 array following the manufacturer’s instructions. Slides were washed and scanned on an Illumina iScan scanner.

#### Bioinformatics

All bioinformatics analysis was carried out on the a high performance computing service. For the amplicon resequencing, FASTQ files were trimmed (Trimgalore) and aligned to the hg19 reference genome using a standard GATK pipeline using the BWA aligner (22). Mutation calling was performed using FreeBayes(23). For the exome sequencing analysis, FASTQ files were trimmed and aligned to the hg19 reference genome using an exome sequencing pipeline Isaac v4 aligner (24), Manta SV caller (25) and Canvas CNV(26) caller. Enrichment was determined by comparison to the Illumina TruSeq exome v1.2 BED file. Mutation calling was performed using Strelka 2 (27) in tumour-normal subtraction mode. For both amplicon sequencing and exome analysis, significantly mutated genes were identified using Intogen(28), MutSigCV (29) and dNdScv (30). Mutational signatures were estimated using the MutationalPatterns (31) R package. Tumour mutational burden was estimating by annotation of the combined, overlapping variant calls from Mutect/Strelka2 to filter to non-synonymous variants (nsV). NsV were then divided by the TruSeq Exome panel size (45.3Mb) to give a figure in mutations/mb.

For structural variant (SV) calls, Manta was used in tumour/normal mode on exome sequencing data and lists of structural variants outputted to VCF file. For copy number calls (CNV), Canvas was used in tumour normal mode and calls outputted as VCF files.

Clonality was determined by running superFreq (32), a cancer specific tumour exome caller and river plots were producing from this package. For neoantigen calls, tumour-normal subtracted VCF files produced with Strelka, GT fields were added via conversion of the SGT field (using a custom script), annotated with the variant effect predictor (VEP), filtered for indels and then analysed using PVacTools against MHC Class I binding predictions (33). Neoantigens with a “best” median IC of < 50nmol.L-1 were counted as binding neoantigens. HLA typing for each patient was determined with HLA-LA on germline exome sequencing data (34).

For methylation, IDAT files were imported into R/Bioconductor and analysed on the ChAMP pipeline (35) using standard settings. iDAT files were imported, standardised to beta methylation values, then underwent SWAN normalisation. Normalised beta-values underwent differential methylation analysis using limma, DMR were called using dmrLasso. Pathway analyses were conducted in GProfiler2 (36).

## RESULTS

### Samples

In the pre-treatment group, 7 biopsies were only available in isolation (because of selecting complete pathological response). In the post-treatment group, there were 24 samples that had had poor response to chemoradiotherapy as defined by a low histological tumour regression grade.

### Sequencing metrics

In total 31 samples successfully underwent amplicon sequencing and 31 samples underwent whole exome sequencing. For the amplicon sequencing average read depth was 1020x (range 357-5210x). For the whole exome sequencing the average read depth was 99x for tumour samples and 55x for control normal. For methylation arrays all samples hybridised successfully.

### Mutational profiles

In the amplicon sequencing group, the top significantly mutated genes as defined by MutSigCV were *PIK3CA* (p=2.4⨯10-9, Q-4.54⨯10-5), *FBXW7* (p=8.68×10-4) and *PTEN* (p=1.02⨯10-3). For dNdScV, the top significantly mutated genes were *AKT1* (p_mis_=2.01⨯10-3), *FBXW7* (p_mis_=5.90⨯10-3), *AMER1* (p_mis_=2.48⨯10-2) and *POLE* (p_mis_=1.32⨯10-2). Gene centric analysis with Intogen demonstrated significant driver mutations in *APC, PIK3CA, PTEN, FBXW7, TP53, SMAD4, FAM123B* and *POLE*. Pathway analysis using Intogen demonstrated 14 pathways (Table 1) being significantly over-represented in the dataset including hsa05210 (Colorectal Cancer, p=1.88⨯10-16, q=1.62⨯10-14), hsa05222 (Small cell lung cancer, p=8.43⨯10-12, q=3.63⨯10-10), and hsa04150 (mTOR pathway, p=4.99⨯10-10, q=1.43⨯10-8)

**Table 1.** Intogen significantly enriched pathways

| ID | Pathway | ZSCORE | PVALUE | QVALUE |
| --- | --- | --- | --- | --- |
| hsa05210 | Colorectal cancer | 8.14 | 1.88E-16 | 1.62E-14 |
| hsa05222 | Small cell lung cancer | 6.73 | 8.43E-12 | 3.63E-10 |
| hsa04150 | mTOR signalling pathway | 6.10 | 4.99E-10 | 1.43E-08 |
| hsa04310 | Wnt signalling pathway | 5.96 | 1.26E-09 | 2.71E-08 |
| hsa05212 | Pancreatic cancer | 5.77 | 3.91E-09 | 6.60E-08 |
| hsa00562 | Inositol phosphate metabolism | 5.71 | 5.37E-09 | 6.60E-08 |
| hsa04070 | Phosphatidylinositol signalling system | 5.71 | 5.37E-09 | 6.60E-08 |
| hsa04115 | p53 signalling pathway | 5.60 | 1.01E-08 | 1.09E-07 |
| hsa05166 | Human T-cell leukemia virus 1 infection | 5.50 | 1.87E-08 | 1.79E-07 |
| hsa05169 | Epstein-Barr virus infection | 4.99 | 2.96E-07 | 2.12E-06 |
| hsa05162 | Measles | 4.99 | 2.96E-07 | 2.12E-06 |
| hsa04210 | Apoptosis | 4.99 | 2.96E-07 | 2.12E-06 |
| hsa05217 | Basal cell carcinoma | 4.66 | 1.56E-06 | 1.03E-05 |
| hsa04120 | Ubiquitin mediated proteolysis | 4.59 | 2.16E-06 | 1.33E-05 |

For the exome sequencing data, analysis by MutSigCV revealed 1,412 genes that were significantly mutated (supplementary tables). The top genes were *HIVEP3, HS6ST3, KIAA1671, LRRC4C* and *ROBO2*. Pathway analysis by GProfiler demonstrated no enrichment of any KEGG pathways. Analysis by Intogen revealed demonstrated 1620 genes significantly mutated (supplementary tables as determined either by OncoDriveClust or OncoDriveFM. The top five genes were *CDC27, CTBP2, IGSF3, PABPC3* and *ZNF432*. Pathway analysis with Intogen demonstrated over-representation of focal adhesion (hsa04510, p=2.25⨯10-135, q=5.82⨯10-133), which contains within it the MAPK/PIK3K and Wnt signalling pathways; as the top rated pathway (supplementary tables). Analysis by dNdScV demonstrated 209 genes significantly mutated (p<0.05), the top five of which were *ZNF717, MUC3A, APC, OR4C5* and *KRAS*. Pathway analysis of these genes by g: Profiler showed enrichment for Wnt signalling (p=4.36×10-2) and WikiPathways WP4258: LncRNA involvement in canonical Wnt signalling and colorectal cancer (p=3.91⨯10-3) and WP710: DNA Damage Response (only ATM dependent, p=1.02⨯10-2). Overlap of all three algorithms was plotted with Venny, there were seven genes that overlapped between all algorithms - *ZFHX3, ROR2, ARID1A, LYST, RPS6KA1, TCF20*, and *DHX37*.

Mutational signature analysis was performed on the exome sequencing samples. The top ranked signatures were signatures three, five and thirty. Signature three is due to defective base repair due to faulty homologous recombination, signature five is due to the effects of transcription coupled nucleotide repair and signature 30 is due to a defect in base-excision repair due to inactivating mutations in NTHL1 (supplementary tables).

Measurement of tumour mutational burden in the exome sequencing dataset demonstrated that in pre-treatment samples that responded to radiotherapy the TMB was consistently high (median = 38.28 muts/mb, range 15.93-86.95) whereas in the non-responding dataset it was significantly lower (median 7.27 muts/mb, range 3.08-47.3, p=0.02 Wilcoxon, Figure 1). Interestingly, in samples where a pre-treatment biopsy was available with a post-treatment specimen, TMB dropped to very low levels (median 5.89, range 1.08-8.65, Figure 1) in the post-treatment specimen.

**Figure 1:**
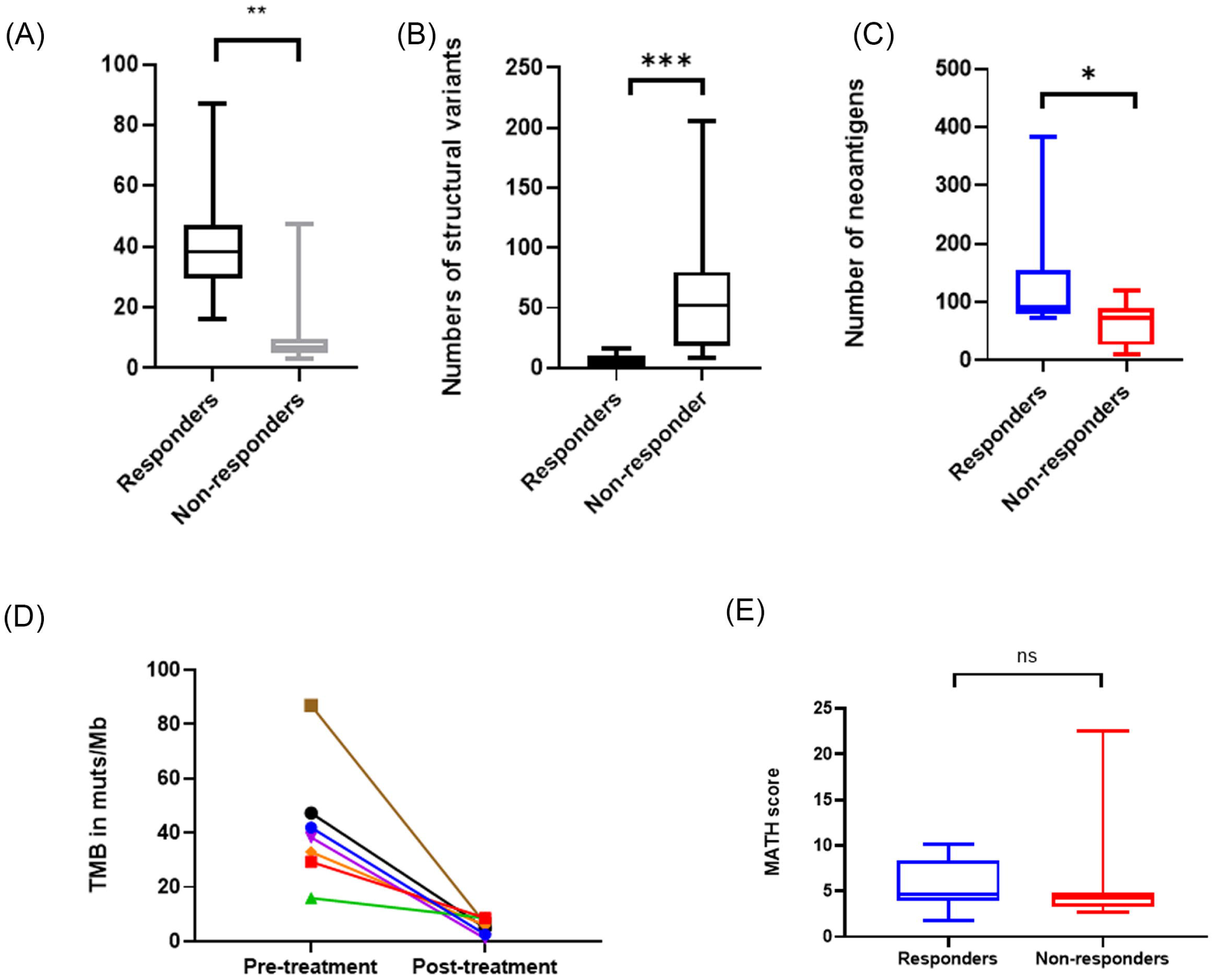
Differences in genomic characteristics between responders and non-responders. A – Tumour mutational burden in mutations/megabase; B-Structural variants per sample; (C) Numbers of neoantigens per sample; (D) Change in Tumour mutational burden in paired samples pre and post response; (E) MATH score per sample

### Structural variants and copy number variation

Copy number estimation was performed on all tumour: normal exome pairs successfully (Figure 2). Four samples showed an extremely complex pattern of copy number gain and loss, which corresponded with complete pathological response to chemoradiotherapy. In the other samples, recurrent copy number alterations were seen at chr12:9660001-9680000, which is an intergenic region that contains a high level of SINE/LINE and may represent an artefact. Otherwise no recurrent copy number alterations were observed.

**Figure 2:**
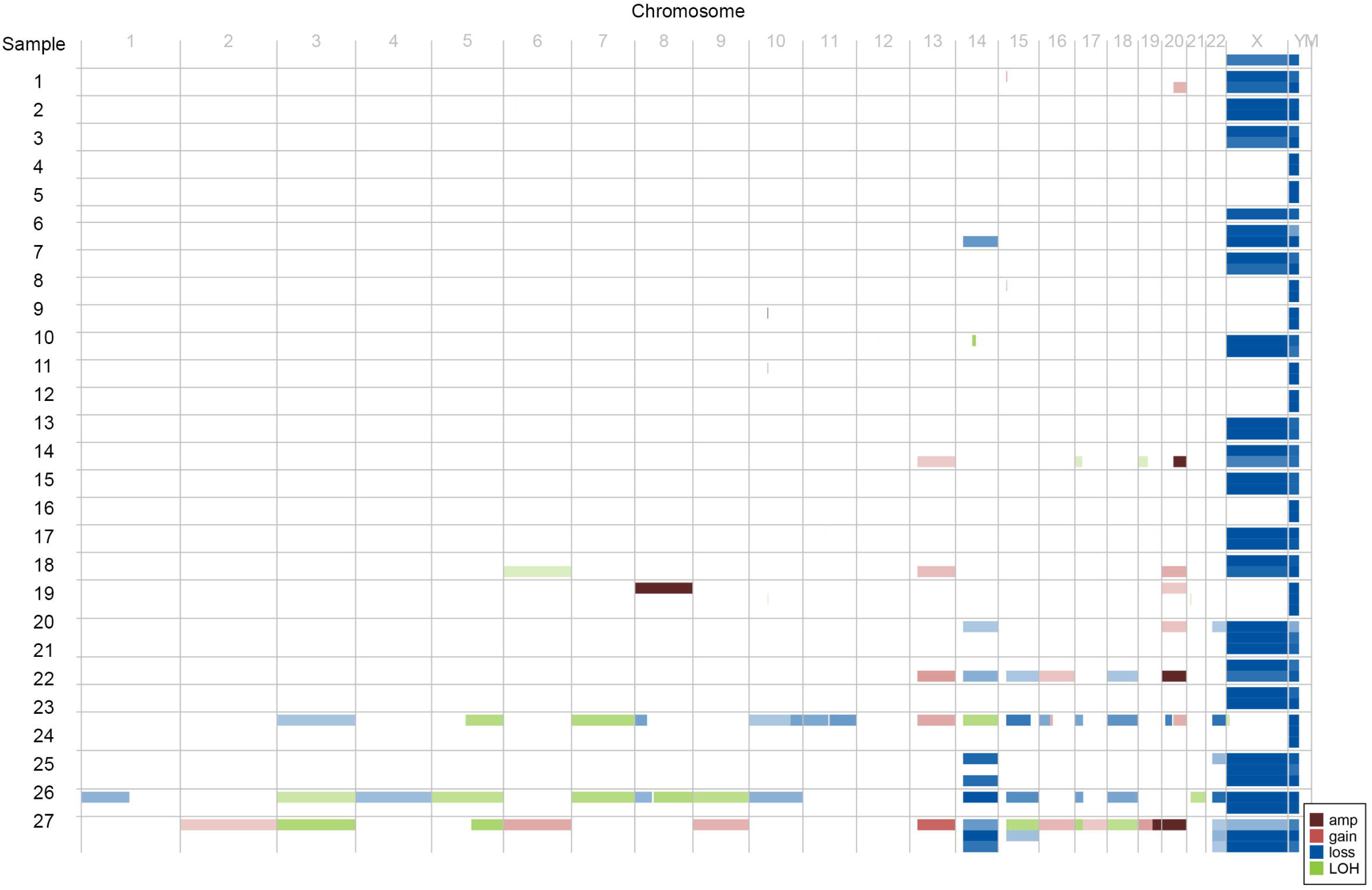
Copy number calls across cohort – ID on Y-axis; Chromsome on X-axis; Amplification in dark red; Gain in light red; Loss in bolue; LoH in green.

Structural variant calling was performed on all tumour: normal exome pairs successfully. The median number of structural variants in responders was 4 (IQR 0-10) vs. 52 in non-responders (IQR 18-80, p<0.001, figure X).

### Clonal evolution analysis

In the seven samples (out of twenty four) where triplet normal, pre-treatment biopsy and post-treatment specimen were available, clonal evolution analysis with superFreq was carried out. This revealed that subsequent to radiation therapy there was an increase in clonal diversity (Figure 4) across all samples with pre-treatment biopsies suggesting where residual tumour remained after radiation therapy this expanded in clonal diversity as a direct result of the selection pressure of radiotherapy. We calculated MATH score, a measure of tumour heterogeneity for all samples, finding that there was a median MATH score of 4.7 in non-responders and 4.2 in responders (Mann Whitney p=0.5036).

**Figure 3:**
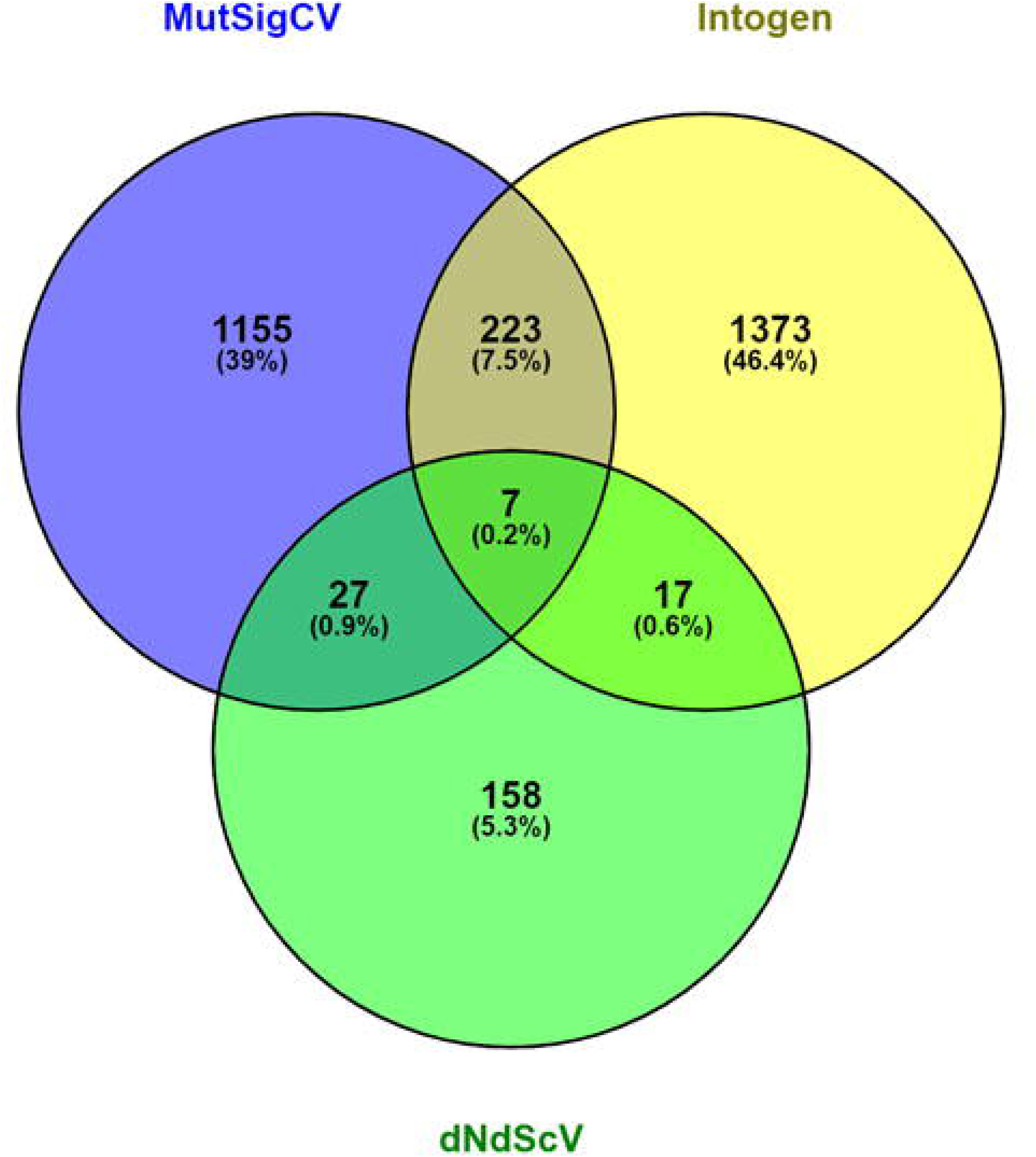
Venn plot of overlaps of callers between three different significant mutation callers

**Figure 4:**
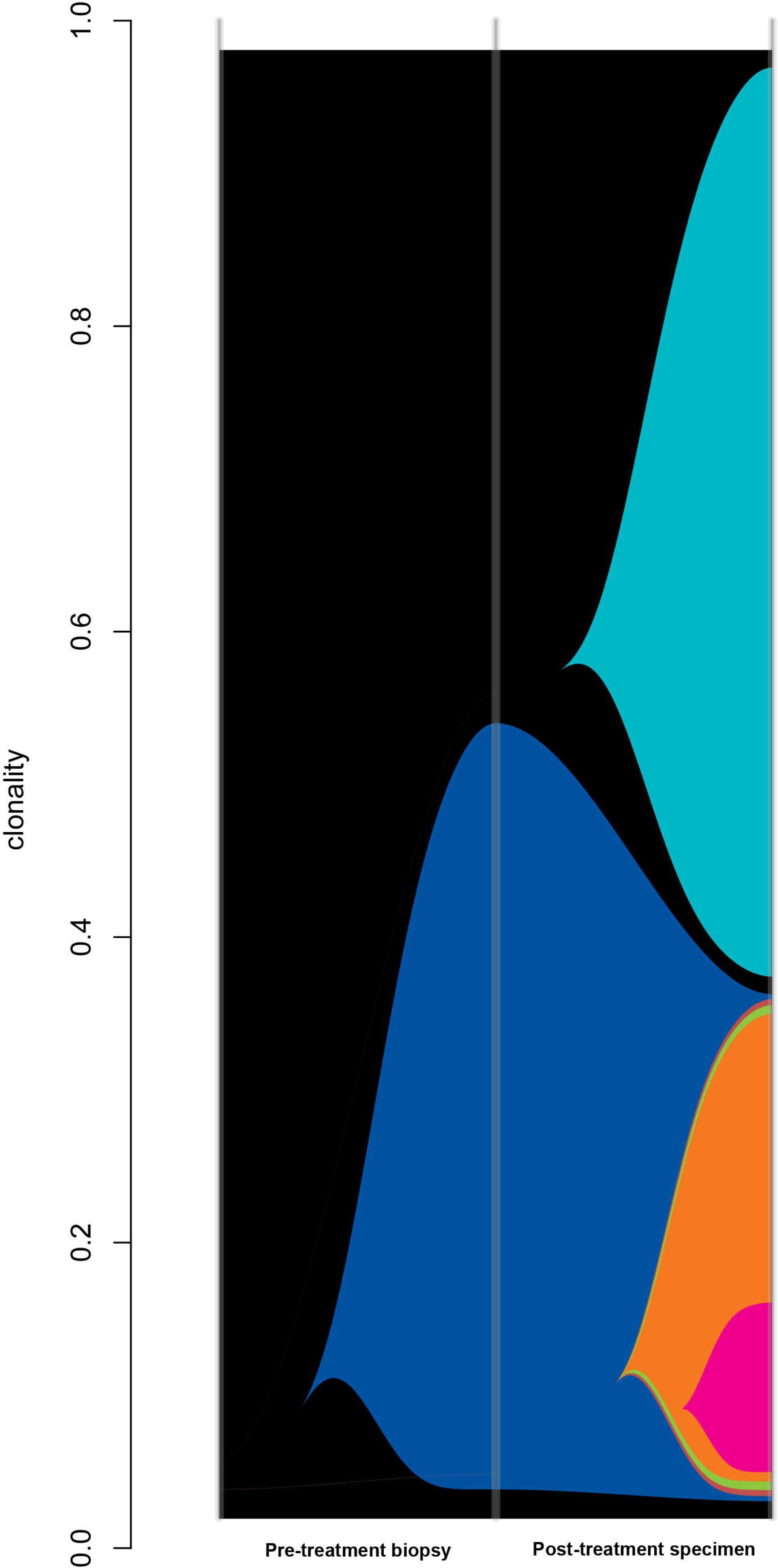
Example river plot of poorly responding cancer; Pre-treatment biopsy demonstrates one clone in sample (clonality 0.57); post treatment specimen shows multiple (five more) clones evolving as a result of radiotherapy selection pressure.

### Methylation analysis

Analysis of the methylome of the tumours undergoing a good response demonstrated 1853 differentially methylated positions, the top ranked probe was cg06982190 (adjusted P-value, 6.21⨯10-15, BF=35.7, deltaBeta=0.33, chr7:27199661-27200960) which tags the CpG island immediately upstream of the *HOXA9* gene. Go:Profiler analysis of these differentially methylated genes demonstrated that the WikiPathways WP306 (Focal adhesion p=1.16⨯10-3), WP185 (Integrin mediated focal adhesion p=2.42×10-3) and WP3932 (PI3K-Akt-MTOR pathway p=3.31⨯10-2) were enriched. Pathway WP306 contains the PI3K/Akt/MTOR signalling genes as well as WP3932 and represents a general enrichment of this pathway.

Analysis of differentially methylated regions demonstrated the top DMR was in the region of chr6:30094947-30095802 downstream of the TRIM40 gene (p-value 0, p-value area=1.67×10-4). HOMER annotation of top 100 DMRs and then analysis by g:Profiler revealed only enrichment in the Human Phenotype pathway HP0000104 Renal Agenesis (p=2.82×10-2) and the GO:MF term Class II MHC binding (p=1.77⨯10-2). Differential block analysis revealed that the top differential methylated block was located at chr1:96420019-96881853 (p_valuearea_=0.03) which is within 50kb of LINC01787. In total, 1795 blocks were differentially methylated, and analysis with g:Profiler demonstrated enrichment in the Axon guidance pathway (KEGG:04360, p=1.94×10-3) and RAP1 signalling pathway (KEGG:04015, p=4.81⨯10-2) as well as the microRNA hsa-miR-21-5p (p=8.36⨯10-9).

### Neoantigen prediction

Neoantigen prediction using pVacTools revealed a median of 78 neoantigens per sample (range 9-383). The hypermutant samples unsurprisingly had the highest neoantigen load as well as the best response to radiotherapy, with a median 91 neoantigens in the responder group and 78 in the non-responder group (Mann-Whitney p=0.0342).

### Immunohistochemical analysis of DSB pathway and mismatch repair deficiency

Semi-quantititative analysis using QuPath was carried out on white light DAB stained images of the expression of yH2AX, Ku70/80, ATM, MLH1, MSH2 and MSH6 respectively.

For the expression of DNA repair genes, significant differential expression was seen (Table 2) in yH2AX, Ku70/80, ATM, MLH1 and MSH6 expression when compared to response/no response status. However, especially for the mismatch repair proteins, despite there being statistically significant differences, these were only a few percentage points different and little complete loss of expression was seen.

**Table 2:** Expression of DNA repair proteins between responders and non-responders

| Antibody | Mean expression non-responders (95% CI), % | Mean Expression responders (95% CI), % | p-value |
| --- | --- | --- | --- |
| yH2AX | 78.2 (73.4-82.9) | 89.8 (83.9-95.6) | 0.0069 |
| ATM | 69.2 (63.4-74.8) | 80.4 (72.2-88.6) | 0.0430 |
| Ku70/80 | 28.3 (23.4-33.1) | 88.3 (84.1-92.5) | <0.0001 |
| MLH1 | 70.2 (65.9-74.6) | 83.3 (77.9-88.6) | 0.0026 |
| MSH2 | 33.3 (28.8-37.9) | 41.7 (30.9-52.5) | 0.1032 |
| MSH6 | 76.1 (72.4-79.9) | 88.2 (84.7-91.6) | 0.0007 |

## CONCLUSIONS

In this study, we have demonstrated a number of interesting findings comparing pre-treatment biopsies and post treatment specimens in patients undergoing (nCRT) for rectal cancer.

The most important finding is that in patients achieving a complete or near complete response to nCRT is that the pre-treatment tumour has a high tumour mutational burden (37). This is because hypermutation within the tumour due to an intrinsic DNA repair defect and leads to increased presentation of neoantigens because of indel and frameshift mutations (38). This increases immunovisibility and we hypothesise that the increased immunovisibility, coupled with activation of the immune system by irradiation, causes cGAS/STING activation which has been shown to lead to a Type I interferon response (39) leading to migration of immune cells and enhanced regression. Also, it suggests that these patients may benefit from anti-PD1/PD-L1 or anti-CTLA4 immunotherapy as high TMB (defined as > 10 muts/mb) has been shown to be correlated with responsiveness to these agents(1). Another intriguing possibility is that genotoxic therapy, coupled with radiation therapy could be delivered as part of neoadjuvant treatment in the tumour, possibly increasing neoantigen burden and making more patients suitable for immunotherapy(40). Our sample size of pathological complete responders is relatively small, however we deliberately chose tumour regression where absolutely no cells remained, which is a very rare phenomenon, compared to the phenomenon of “minimal residual disease”, but we believed that this would give a stronger biological signal.

We have also demonstrated that there is enrichment for mutations in the mTOR/AKT signalling pathways, specifically *PIK3CA* but also as a general trend towards mutations and epigenetic changes in this pathway. *PIK3CA* is a member of the mTOR signalling pathway and makes up the alpha subunit of the PI3K protein. mTOR signalling has previously been highlighted as being of possible relevance in radiosensitivity (41) as a cellular marker of stress in the tumours of patients undergoing chemoradiotherapy. *PIK3CA* signals through the mTORC1/mTORC2 and exerts its downstream effect on AKT (42). Targeted agents for *PIK3CA* (apitosilib (43)), mTORC1/2 (visusertib (44)) and AKT (MK2206 (45)) exist and are at various stages of clinical development. We suggest that these agents may be utilised as part of a neoadjuvant therapy strategy in order to increase response rates.

*FBXW7*, a gene previously implicated in cell cycle control by ubiquitination? of cyclin-E1 (46) was also significantly enriched for mutations within it in this study. Zhang et al demonstrated that *FBXW7* also had a role in non-homologous end joining (47, 48), being a binding partner in the complex that repairs damage caused by double strand breaks. Mutations in *FBXW7* may affect its ability to participate in NHEJ and therefore increase radiosensitivity. Mutational signature analysis of the exome sequencing dataset also showed multiple mutational signatures consistent with enrichment in DNA repair, specifically faulty homologous recombination (HR) and nucleotide excision repair.

Tumour heterogeneity is a significant problem across all tumours due to drivers that may cause a differential response to therapy because of mutational clonality. Our results show, unsurprisingly, that treatment of samples with neoadjuvant chemoradiotherapy causes an increase in clonal diversity, which may be a driver in the phenomenon of radiation resistance. However, we did not demonstrate any difference between responders and non-responders in terms of MATH score as a measure of tumour heterogeneity, unlike other papers, however the accuracy of MATH score as a measurement of heterogeneity has been called into question because of differences in variant callers (49).

In the epigenetic analysis, the microRNA pathway Has-mir-21-5p was significantly differentially methylated. This microRNA is of significant interest (50) in response to therapy as it represents a key driver in the cellular response to hypoxia. The immunohistochemical analysis of tumour samples showed increased yH2AX and ATM expression was associated with response, as well as increased expression of the mismatch repair proteins MLH1 & MSH6. We found this puzzling, as we would have expected the opposite to be true, i.e. loss of expression was needed for response, especially in ATM and yH2AX. However we hypothesise that these samples may have had regions of loss of normal response and these disappeared as a consequence of response to radiotherapy.

Therefore we suggest that based on these findings, a number of factors contribute to the response to neoadjuvant chemoradiotherapy: hypermutation leading to increased neoantigen presentation; enrichment in defects in the mTOR signalling pathway; hypoxia regulated by miR-21-5p and an increase in clonal diversity. Our findings agree with those of Akiyoshi et al (1) in that increase in neoantigen diversity correlated with response. Kamran (13) however, found no clear molecular defects that predisposed to radioresistance. This could be due to the fact that their analysis was geared towards pathways of treatment resistance rather than those that lead to radiosensitivity.

Clearly, the best way to understand these defects and investigate them further would be to build a cellular model of rectal cancer in order to modulate these pathways in order to measure responsiveness (51). Current cell lines have a bias towards their micro environment and although provide reasonable models of single pathway alterations lack the fidelity to measure therapy response when modulated. The ideal model would be an organoid based rectal cancer therapy model as this provides both the 3D structure (enabling cell/cell communication a more representative element of intra-tumoural hypoxia) and more accurate response to therapy, as well as the ability to evolve and resist therapy.

Our findings suggest a number of new therapeutic avenues for increasing responsiveness to chemoradiotherapy in rectal cancer. We plan to further study these in complex 3D models such as organoids in order to understand whether they will increase response rates when appropriately targeted.

## Data Availability

Sequencing data will be uploaded to EGA on acceptance of the manuscript

## Notes

Funding: This study was supported by grants from the Academy of Medical Sciences and the Wellcome Trust (ref 102732/Z/13/Z). ADB is currently supported by a Cancer Research UK Advanced Clinician Scientist award (ref C31641/A23923)

Conflicts of interest: The authors declare no conflict of interest

### Competing Interest Statement

The authors have declared no competing interest.

### Funding Statement

This study was supported by grants from the Academy of Medical Sciences and the Wellcome Trust (ref 102732/Z/13/Z). ADB is currently supported by a Cancer Research UK Advanced Clinician Scientist award (ref C31641/A23923)

